# A Hidden Structural Variation in a Known IRD Gene: A Cautionary Tale of Two New Disease Candidate Genes

**DOI:** 10.1101/2021.10.29.21265657

**Authors:** Hilary A. Scott, Anna Larson, Shi Song Rong, Sudeep Mehrotra, Rossano Butcher, Katherine R. Chao, Janey L. Wiggs, Emily M. Place, Eric A. Pierce, Kinga M. Bujakowska

## Abstract

Rod cone dystrophy (RCD), also known as retinitis pigmentosa, is an inherited condition leading to vision loss, affecting 1/3500 people. Over 270 genes are known to be implicated in the inherited retinal degenerations (IRDs), yet genetic diagnosis for ∼30% IRD of patients remains elusive despite advances in sequencing technologies. The goal of this study was to determine the genetic causality in a family with Rod-cone dystrophy (RCD). Family members were given a full ophthalmic exam at the Retinal Service at MEE and consented to genetic testing. Whole exome sequencing (WES) was performed and variants of interest were Sanger validated. Functional assays were conducted in zebrafish along with splicing assays in relevant cell lines to determine the impact on retinal function.

WES identified variants in two potential candidate genes that segregated with disease: *GNL3* (G Protein Nucleolar 3) c.1187+3A>C and c.1568-8C>A; and *PDE4DIP* (Phosphodiester 4D Interacting Protein) c.3868G>A (p.Glu1290Lys) and c.4603G>A (p.Ala1535Thr). Both genes were promising candidates based on their retinal involvement (development and interactions with IRD-associated proteins), however the functional assays did not validate either gene. Subsequent WES reanalysis with an updated bioinformatics pipeline and widened search parameters led to the detection of a 94bp duplication in *PRPF31* (pre-mRNA Processing Factor 31) c.73_266dup (p.Asp56GlyfsTer33) as the causal variant.

Our study demonstrates the importance of thorough functional characterization of new disease candidate genes, and the value of reanalyzing NGS sequence data, which in our case led to identification of a hidden pathogenic variant in a known IRD gene.

## INTRODUCTION

Rod-cone degeneration (RCD) also known as retinitis pigmentosa (RP) is the most prevalent type of inherited retinal disease (IRD) which causes progressive vision loss affecting 1/3500 people worldwide (Haim, 2002). IRDs can be inherited as autosomal recessive, autosomal dominant, chromosome X-linked and mitochondrial traits. Genetic diagnosis may additionally be complicated due to the genetic heterogeneity of IRDs with a wide spectrum of clinical phenotypes attributed to pathogenic variants in over 270 genes (RetNet - Retinal Information Network (uth.edu). Targeted exome panels that include known IRD genes as well as whole exome sequencing are the standard approach for determining genetic diagnosis. These are largely focused on identifying single nucleotide variants (SNV), small indels and increasingly large copy number variations, allowing for the identification of causal mutations in ∼60% of cases (Carss et al., 2017; Consugar et al., 2014; Ellingford et al., 2018). Current gene-based therapies require accurate molecular diagnosis which provides an important incentive for determining the cause of disease in the remaining patients (Bainbridge et al., 2008; MacLaren et al., 2014; Maguire et al., 2008). A potential source of this missing causality could be found in novel candidate IRD genes. Although rare, new IRD disease genes have been discovered within the last few years for example: *ARSG, CLCC1, POMGNT1, REEP6, AHR, IFT172* (Arno et al., 2016; Bujakowska et al., 2015; Khateb et al., 2018; Li et al., 2018; Xu et al., 2016; Zhou et al., 2018). Co-segregation of the variant with the ocular phenotype in large pedigrees is often the only indicator of a novel gene initially. In order to classify the variant as causal, first a gene-disease association must be established which requires further genetic and experimental investigation. Candidate genes harboring rare variants can be prioritized based on a previously characterized roles in retinal development or function, interaction of the encoded protein with known IRD-associated proteins (Audo et al., 2009; Zeitz et al., 2006; Zeitz et al., 2005), *in silico* pathogenicity prediction scores (Ionita-Laza et al., 2016; Jaganathan et al., 2019; Kircher et al., 2014; Schwarz et al., 2014; Siepel et al., 2005), and thorough validation with model organisms to characterize the impact on retinal function (Bujakowska et al., 2015).

In this study we applied the above criteria to search for novel candidate gene in a large family with three affected siblings diagnosed with RCD. Two strong candidate genes were found to co-segregate with disease. *G protein nucleolar 3* (*GNL3*) and *Phosphodiesterase 4D interacting protein* (*PDE4DIP*) both encode proteins that are involved with retinal development or known to interact with IRD-associated proteins (Kawashima et al., 2009; Overlack et al., 2011; Paridaen et al., 2011). Possible pathogenicity of the identified variants in both genes was studied in cell and zebrafish models, however after exhaustive investigation, both candidate genes were excluded as the cause of disease. Ultimately the cause of disease was determined to be a difficult-to-detect structural variation in a known IRD gene, *PRPF31 (Vithana et al., 2001)*. In this case, partial penetrance displayed in the unaffected mother and sibling further complicated the genetic analysis due to a misleading inheritance pattern that was actually determined to be autosomal dominant with haploinsufficiency (Abu-Safieh et al., 2006; Vithana et al., 2003). Our study demonstrates the importance of a thorough validation of new candidate genes and the value of sequence data re-analysis.

## RESULTS

A female in her thirties (OGI842-1649) was seen in the IRD clinic at MEE with RCD. Family history revealed that five out of seven children in the family suffered from visual impairment, with no other history of retinal degeneration in the family (Fig. 1A). Subject OGI842-1649 had decreased visual acuity, restricted visual fields and severely reduced cone responses in full-field ERG (Table 1). Her fundus images showed characteristic signs of rod-cone degeneration including peripheral bone spicule pigmentation, vessel attenuation and abnormal granularity in the macula RPE (Fig. 1B). Both parents were evaluated and showed no signs of retinal abnormalities and full-field ERGs were within normal ranges (Fig 1B, Table 1).

**Table 1.**
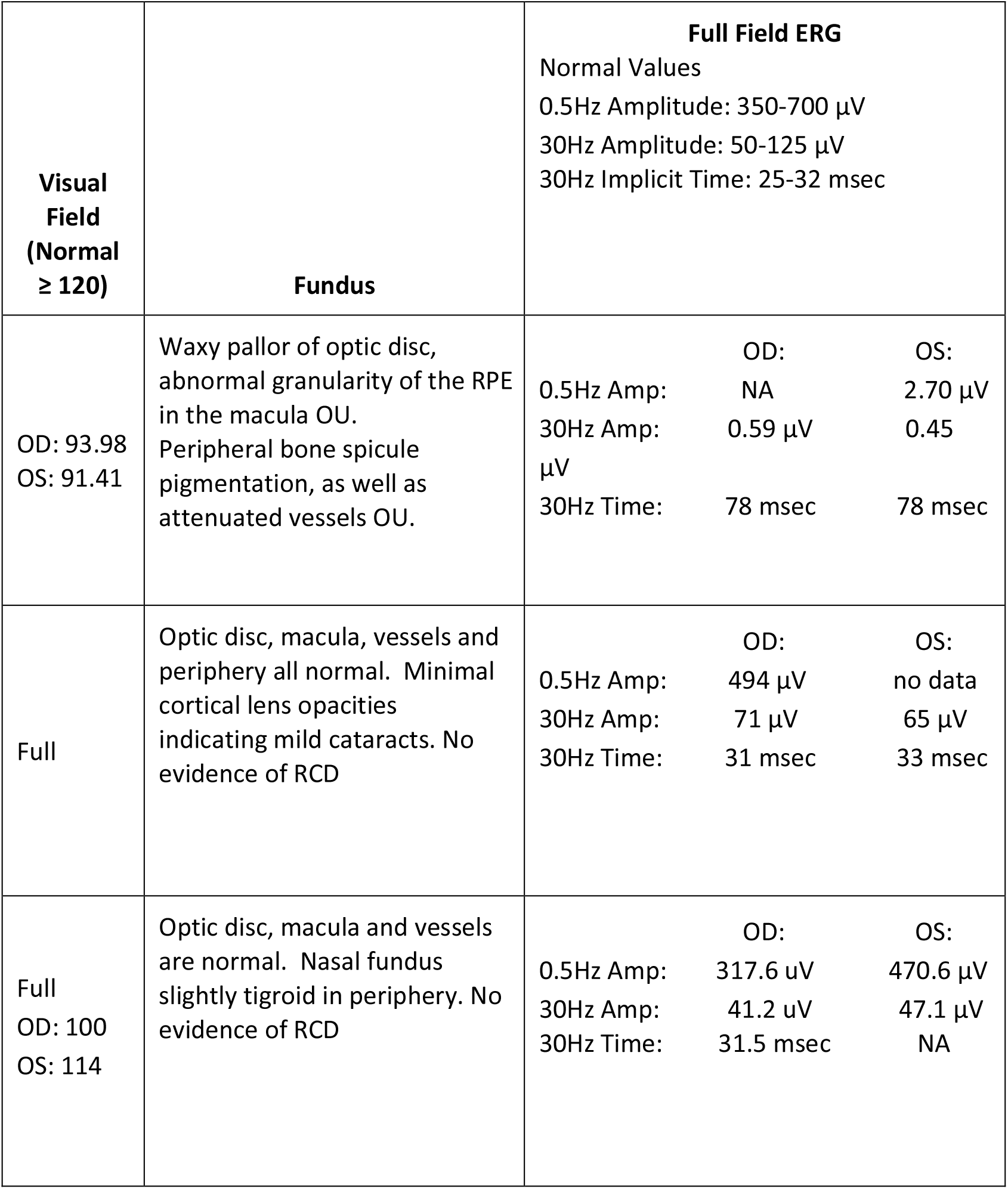
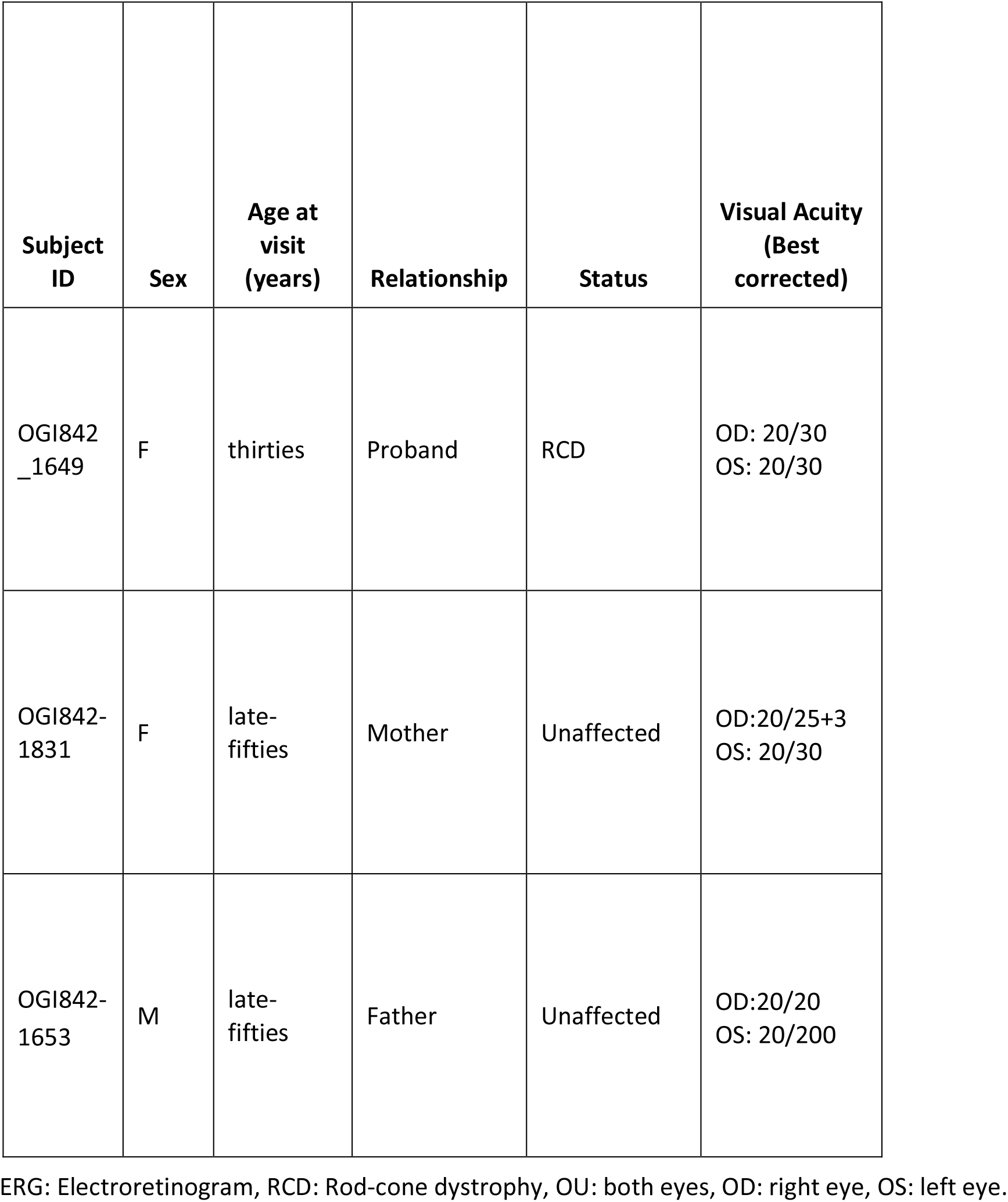
Clinical Phenotypes of Proband and Parents.

**Figure 1.**
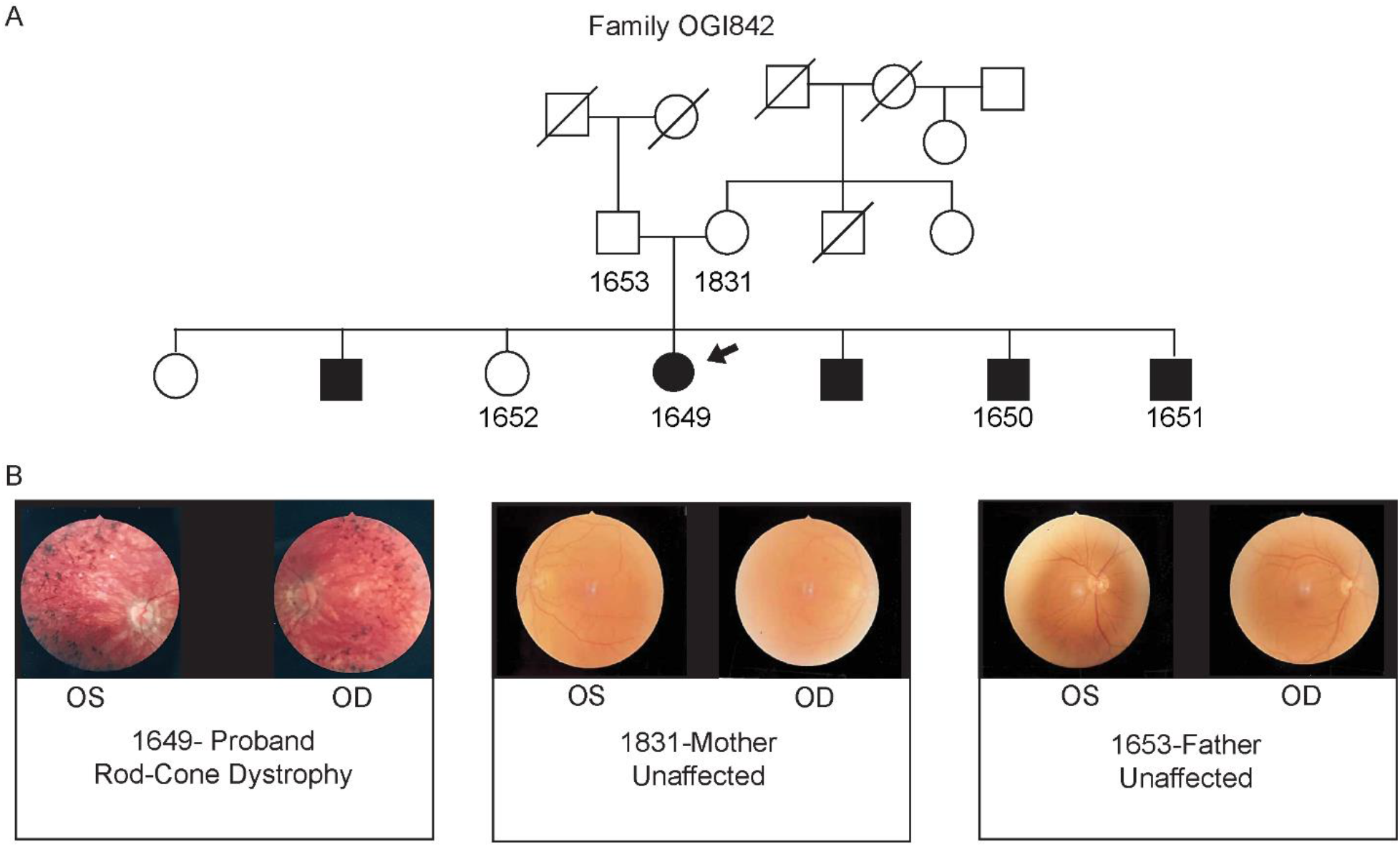
Pedigree and Phenotypic details for Family OGI842. (A) Multigenerational pedigree of OGI842 family. Affected are shown with darkened symbols, proband denoted with arrow, and all subjects that were sequenced have a number designation. (B) Fundus images of the proband (OGI1649) in her early thirties, unaffected mother (OGI1831) in her late fifties, and unaffected father (OGI1653) in his late fifties.

Molecular analysis was performed on the parents (OGI842-1653, OGI842-1831), and four siblings for whom DNA was available (OGI842-1649, OGI842-1650, OGI842-1651, and OGI842-1652). Potential causal variants obtained from exome sequencing were filtered based on minor allele frequency (MAF) ≤0.1%. WES analysis identified rare bi-allelic variants in two candidate genes, *G protein nucleolar 3* (*GNL3*) with c.1187+3A>C and c.1568-8C>A splice region variants (Fig.2A) and *Phosphodiesterase 4D interacting protein* (*PDE4DIP*) with two missense variants (c.3868G>A, p.Glu1290Lys and c.4603G>A, p.Ala1535Thr) (Fig. 3A). Variants in both candidate genes were rare or absent in gnomAD database and were selected for further analysis based on a combination of evolutionary conservation, aberrant splicing, and pathogenicity predictions (Table 2) (Karczewski et al., 2019).

**Table 2.** Variants identified in Whole exome sequencing.

| Gene | cDNA change | Consequence | CADD | Mutation Taster | Splice AI | gnomAD | PhastCon | Eigen | ClinVar | ACMG |
| --- | --- | --- | --- | --- | --- | --- | --- | --- | --- | --- |
| *GNL3* | c.1187+3A>C | Exon skipping | 14 | Disease causing | 0.75 | Absent | 0.999 | 18 | NR | **LP**, Likely pathogenic<br>PVS1, PS3, PM2, PP1, PP3, BP5 |
| *GNL3* | c.1568-8C>A | NA | 7.4 | Disease causing | 0.15 | Absent | 0.001 | 8.1 | NR | **VUS**, Uncertain significance, PM2, PM3, PP3, PP4, BS3, BP5 |
| *PDE4DIP* | c.3868G>A | p.Glu1290Lys | 32 | Disease causing | 0.0 | 1.32x10<sup>-5</sup> | 1.00 | 6.9 | NR | **LB**, Likely benign, PM2, PP3, PP4, BS3, BP5 |
| *PDE4DIP* | c.4603G>A | p.Ala1535Thr | 23 | Poly-morphism | 0.0 | $3.281 \times 10^{-4}$ | 0.773 | 1.0 | NR | **LB**, Likely benign, PM2, PP3, PP4, BS3, BP5 |
| *PRPF31* | c.73_166dup | p.Asp56Glyfs Ter33 | NA | Disease causing | NA | Absent* | NA | NA | LP | **P**, Pathogenic PVS1, PM2, PP1, PP3, PP4 |

CADD, Combined Annotation Dependent Depletion; gnomAD, Genome Aggregation Database; PhastCon, PHylogenetic Analysis with Space/Time models; ACMG, American College of Medical Genetics and Genomics; NR, not reported; Absent\* from both gnomAD and gnomAD-SV.

**Figure 2.**
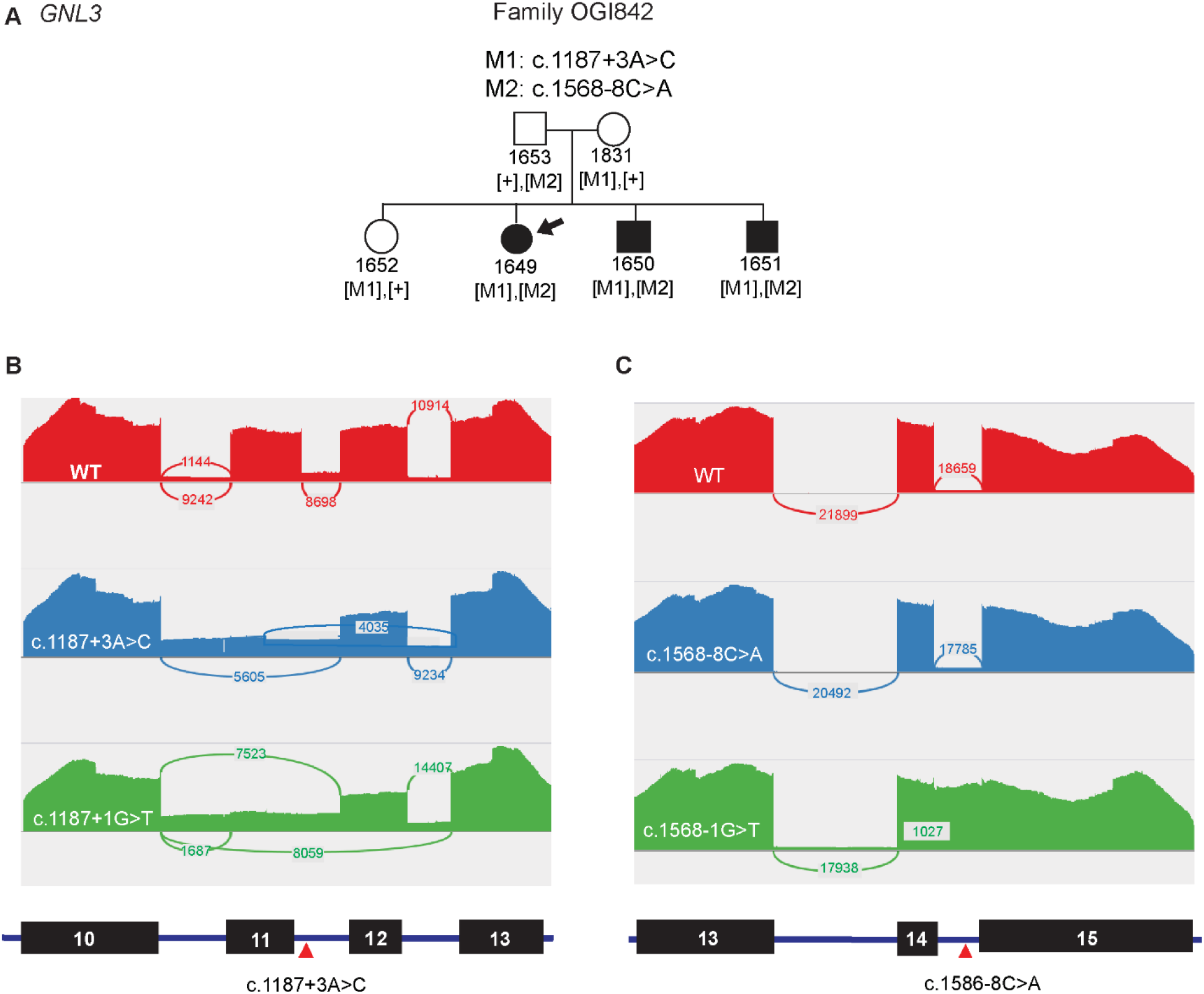
Investigating variants in *GNL3*, a candidate gene for IRD. **(A)** Pedigree analysis confirms segregation of *GNL3* with disease, where filled symbols represent affected individuals and empty symbols represent healthy individuals, circles are females and squares are males. **(B)** NGS analysis of mRNA splicing of the exon10-13 mini gene construct. Read coverage and exon junctions are represented by sashimi plots for the wild type control (red), the c.1187+3A>C variant (blue) and the essential splice site control c.1187+1G>T (green). The corresponding mini gene transcript model with the position of the mutation indicated with a red arrowhead is shown below. Exon 11 skipping (5605 junction reads) and intron 10 retention with partial exon 11 inclusion and exon 12 skipping (4035 reads) are the major effects of the c.1187+3A>C variant. Exon 11 skipping (7523 reads) and exon 11/12 skipping (8059) reads are the major essential in the essential splice site control. Normalizing exon 11 splicing events to adjacent Skipping of exon 11 occurs in 60% of transcripts (5605/9234 read ratio) as compared to wild type 9242/10914, exon 12 skipping occurs 4035/9234. **(C)** Sashimi plots generated from the amplicon sequencing on wild type (red), c.1568-8C>A (blue) and essential splice site mutation c.1568-1G>T (green) across regions mini gene including exons 13-15. No difference is observed in the splicing pattern between the wild type control and the c.1568-8C>A variant. Essential splice site control, c.1568-1G>T, showed complete intron 14 inclusion.

**Figure 3.**
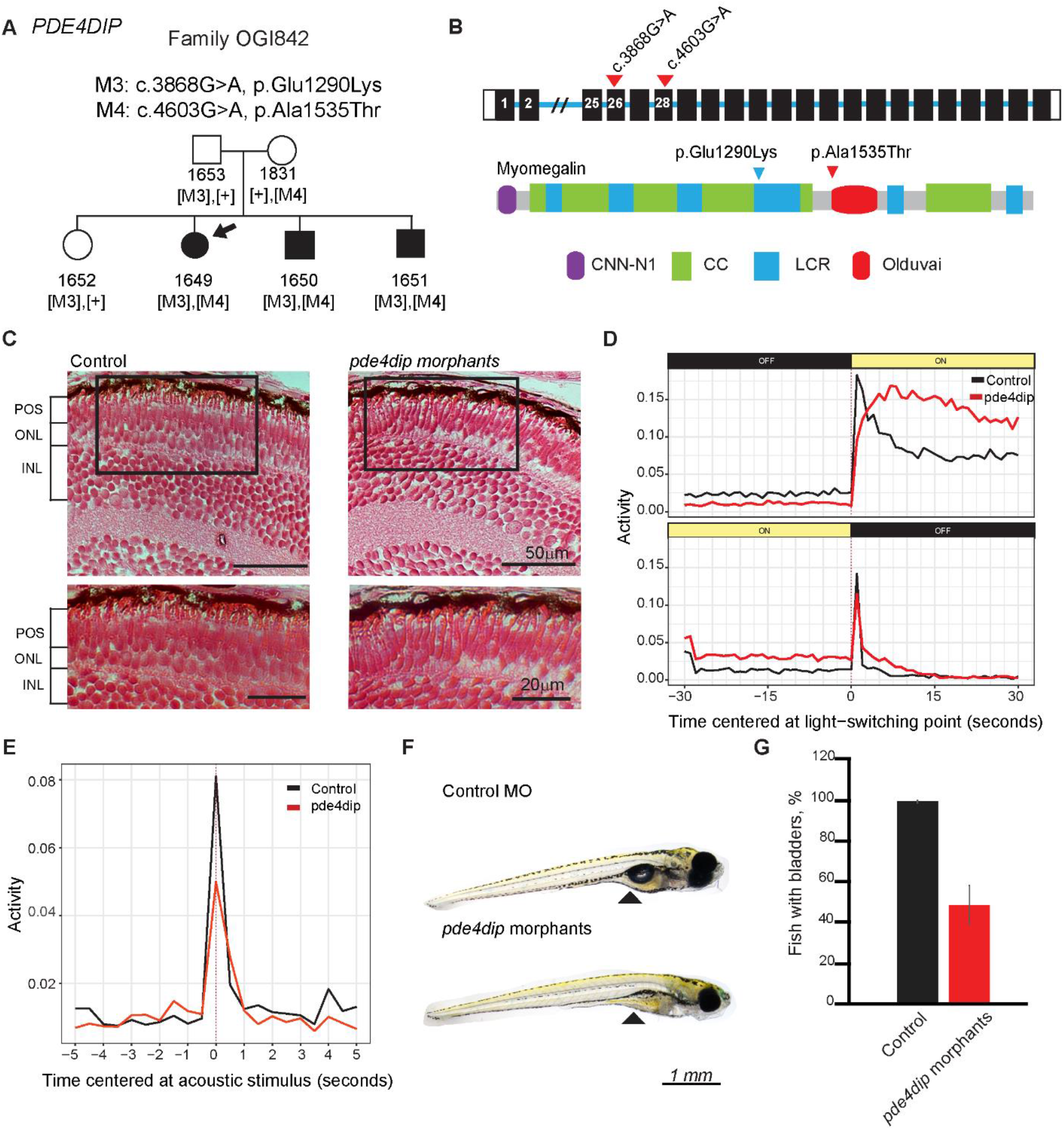
Investigating *PDE4DIP* as a candidate gene for IRD. **(A)** Pedigree analysis confirming segregation of p.Glu1290Lys and p.Ala1535Thr missense variants. **(B)** Schematic of the *PDE4DIP* gene and the encoded protein (Myomegalin, PDB: MYOME_HUMAN (Q5VU43)) with mutations indicated (red arrowheads) in exons 26 and 28. **(C)** Retinal histology of *pde4dip* morphants at 5 days post fertilization (dpf), showing no observable differences in morphology as compared to controls. **(D)** VMR assay showing a significant difference in response to light stimuli in *pde4dip* morphants (red, n=144) and control MO (black, n=143) (Tukey HSD, Light OFF p= 2.73+10^−13^, Light ON p=0.0136). **(E)** AEBR assay showing decreased response in *pde4dip* morphants (red) compared to control fish (black) (n=144, Tukey HSD, p=0.8996) indicating general locomotor abnormalities. **(F)** Zebrafish analyzed at 5 dpf showing absence of gas bladders in *pde4dip* morphants. **(G)** Quantification of the gas bladder presence in *pde4dip* morphants (red) (n=134, 54.48%) versus control fish (black) (n=134, 98.51%) Fisher’s exact test p<0.0001.

*GNL3* encodes G protein nucleolar 3, also known as nucleostemin, which maintains cell proliferation and is necessary for the correct timing of cell cycle exit and differentiation (Tsai and McKay, 2002). *GNL3* is expressed in retinal progenitor cells (Schmitt et al., 2009), and zebrafish *gnl3* mutants were shown to have smaller eyes and delayed retinal cell development (Paridaen et al., 2011). Therefore, we hypothesized that the two splice region variants c.1187+3A>C and c.1568-8C>A in *GNL3* may lead to retinal dysfunction. Both changes were absent in gnomAD and TOPMed. The c.1187+3A>C change, located in intron 11, was predicted to affect splicing by multiple splicing prediction software, while c.1568-8C>A in intron 14 had an elevated Eigen score (Ionita-Laza et al., 2016), and both were predicted to be disease causing by Mutation Taster (Table 2)(Schwarz et al., 2014). Both variants were studied by splicing assays in HEK293T cells. The splicing constructs spanned exons surrounding the studied variants with two controls: a wild type and an essential splice site control (Fig. 2 B, C). Next generation sequencing (NGS) analysis of the midi-gene transcripts revealed skipping of exon 11 and 12 in the c.1187+3A>C variant and essential splice site control (Fig. 2B). However, the second studied variant, c.1568-8C>A, had no effect on splicing (Fig. 2C). This variant was therefore considered to be nonpathogenic. In addition, knocking down *gnl3* with a splice blocking morpholino oligonucleotide (MO) in zebrafish did not show any morphological or functional effects on the eye or the retina (Supplementary material Fig. S1). These findings conclusively dismissed *GNL3* as being the cause of disease in this family.

We therefore turned our attention towards the second candidate gene, *PDE4DIP* whose product, myomegalin, is involved in cAMP-dependent signaling (Verde et al., 2001) and organization of centrosomal and Golgi-derived microtubules (Wang et al., 2014). Isoforms of myomegalin in the retina have been shown to interact with Sans, encoded by *USH1G*, Usher syndrome 1G (Overlack et al., 2011). Loss of *USH1G* causes congenital hearing loss and RCD (Mustapha et al., 2002) thus the implication of *PDE4DIP* playing a role in the Usher interactome made it a promising candidate gene for retinal degeneration (Weil et al., 2003). WES detected two missense variants in *PDE4DIP* (c.3868G>A, p.Glu1290Lys and c.4603G>A, p.Ala1535Thr; NM_001198834.3) that are rare in gnomAD and are highly conserved (Karczewski et al., 2019) (Fig. 3A-B, Table 2).

Zebrafish studies were carried out to determine the effect of *pde4dip* MO knock-down on the retina (Fig.3C-D). Even though no structural retinal abnormalities were observed (Fig. 3E), *pde4dip* morphants showed reduced function in the visual motor response (VMR) (Fig. 3D). To exclude the possibility that the reduced VMR response was a result of other developmental anomalies rather than retinal malfunction, we performed an acoustically evoked behavioral response (AEBR) assay. AEBR measures the motor response after an acoustic stimulus, providing a vision independent control. In the AEBR experiment, *pde4dip* morphants had lowered startle response compared to control fish indicating other vision unrelated developmental defects affecting motor response (Fig.3E). Upon a closer inspection of the fish morphology, we realized that ∼50% of the *pde4dip* morphants showed maldeveloped gas bladders, which indicates a general swimming impediment and explains reduced or lack of startle response upon visual or acoustic stimulation (Fig. 3F). Similar experiments repeated in CRISPR/Cas9 generated *pde4dip* mutant larvae also showed no retinal phenotype. (Supplementary Fig. S4). Given this, we concluded that the *pde4dip* morphant phenotype is not specifically related to vision and that the loss of *pde4dip* in zebrafish and likely other organisms has no effect on retinal development and is unlikely the cause of disease in the studied family.

As both candidate genes were disproven, WES data was reanalyzed. Removing the inheritance and variant quality filters (variant quality score recalibration) (McKenna et al., 2010a) led to the identification of a 94 nucleotide tandem duplication within the first coding exon of Pre-mRNA Processing Factor 31, *PRPF31*, which is expressed in all tissues, including the retina, (Lonsdale et al., 2013) https://www.gtexportal.org/home/gene/PRPF31). The *PRPF31* encoded protein, U4/U6 small nuclear ribonucleoprotein Prp31, facilitates protein-RNA interactions required for spliceosome assembly. The retina may be particularly sensitive to perturbations in splicing as mutations in splicing factors (Tanackovic et al., 2011), including *SNRNP200, PRPF3, PRPF8*, and *PRPF31* among others, are known to cause autosomal dominant rod-cone dystrophy (Chakarova et al., 2002; McKie et al., 2001; Vithana et al., 2001; Zhao et al., 2009). The duplication, c.73_166dup, leads to a frameshift that creates a stop codon, p.Asp56GlyfsTer33 (Fig.4A), likely resulting in nonsense mediated decay of the transcript (Fig. 4B). PCR amplification of the region surrounding the duplication confirmed that all three affected family members (OGI842-1649, OGI842-1650, OGI842-1651) carry this variant, as well as the unaffected sibling (OGI842-1652) and the mother (OGI842-1831), indicating incomplete penetrance (Fig.4C). Sanger sequencing confirmed mapping of the duplication to hg19: chr19:54,621,731-54,621,824 (Fig.4D). The disease mechanism of *PRPF31* is known to be haploinsufficiency and therefore the uncovered heterozygous loss-of-function variant was determined to be the cause of disease in this family, with two asymptomatic mutation carriers. Partial penetrance, a known feature of *PRPF31*-associated RCD (Rivolta et al., 2006; Vithana et al., 2003), explains the initial assumption that disease inheritance in this pedigree was autosomal recessive. The inheritance of the wild type allele was shown to determine disease penetrance (Rivolta et al., 2006; Vithana et al., 2003), which was also seen in the presented family, where the unaffected sibling inherited a different wild type allele from her father than the affected siblings (Fig. 4A).

**Figure 4.**
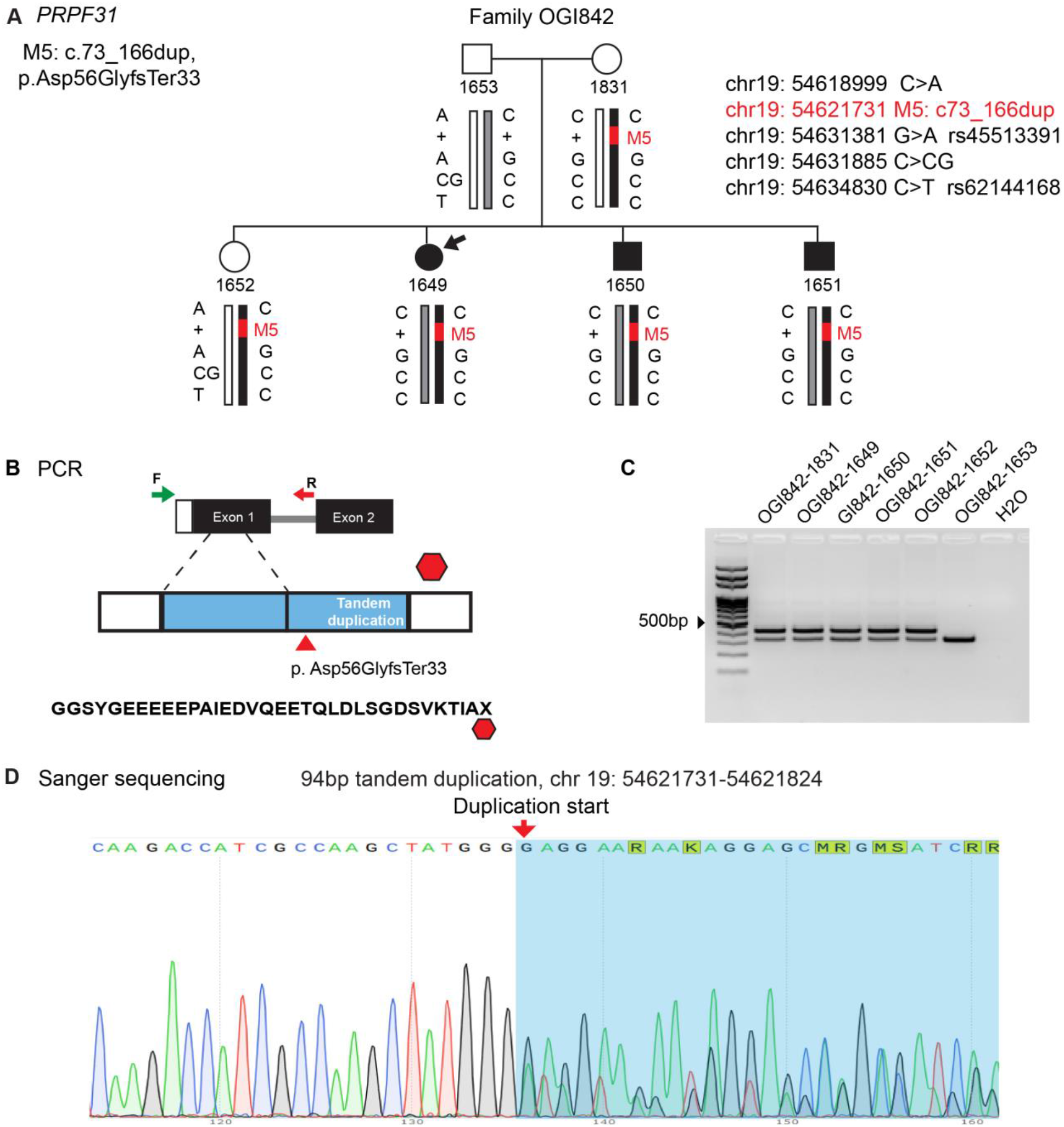
Validation of *PRPF31* duplication. **(A)** Familial segregation of the c.73_166dup, p.Asp56GlyfsTer33 variant (red bar) on maternal allele revealing autosomal dominant inheritance pattern with partial penetrance. Segregation of the paternal wild type allele among siblings with unaffected (white bar) and affected (gray bar). **(B)** A schematic diagram of the first two coding exons of *PRPF31* showing primers used to validate the proposed tandem duplication below. The altered amino acid sequence (p.Asp56GlyfsTer33) of the duplication results in the creation of a stop codon (red octagon). **(C)** PCR amplification of gDNA from all family members showing the heterozygous duplication (two bands) in all but the father (OGI842-1653). The lower band 335bp is expected for wild type allele, the upper band 429bp is expected with the inclusion of the 94bp duplication. **(D)** Chromatogram for Sanger sequencing shows start point for duplication with genomic location hg19 chr19:54621731-54621824.

## DISCUSSION

Our study describes a search for the genetic cause of disease in an RCD family whose inheritance pattern appeared to be autosomal recessive with three affected members. Whole exome sequencing identified rare variants in two candidate genes that segregated with the disease phenotype and met the criteria determined in the outset of the study being highly conserved, rare or absent in genome aggregation databases, predicted to be disease causing and having relevant interactions with known IRD genes. The first candidate gene investigated *GNL3*, also known as nucleostemin, belongs to a family of Nucleolar GTP-binding proteins which includes members *GNL3L* and *GNL2* which has been shown to be expressed in retinal precursors (Kawashima et al., 2009; Schmitt et al., 2009). Zebrafish models show both that *gnl2*^*-/-*^ and *gnl3*^*-/-*^ play a role in retinal neurogenesis with their loss leading to delayed retinal differentiation. Proteins coded by these genes have partly redundant function with *gnl3*^*-/-*^ displaying a milder phenotype with transient abnormalities at early stages of development at 22 hours post fertilization (hpf), which resolve at later time points (Paridaen et al., 2011). However, our results showed no phenotype associated with knock down of *gnl3* using a splice blocking morpholino. Our attempts to employ a translational blocking morpholino that would have arguably been a stronger knock down and more closely comparable to the previous knock out model, resulted in a “monster” phenotype and therefore could not be studied further. The lack of developmental phenotype in our *gnl3* morphants may also be due to the fact that we evaluated the morphant larvae at a later developmental stage, at 5dpf rather than 22 hpf (Paridaen et al., 2011). Ultimately, the lack of phenotype in our zebrafish knockdown, as well as the absence of splicing aberrations in in vitro assays, led us to exclude *GNL3* as the cause of disease in this family. The second gene, *PDE4DIP*, encodes myomegalin, which has been shown to interact with SANS, an Usher interactome protein encoded by *USH1G* (Overlack et al., 2011). In the end, functional assays in zebrafish failed to demonstrate *PDE4DIP* impacted retinal function. Our results thus demonstrate the importance of not only the stringent selection criteria for candidate genes but also of rigorous functional validation to avoid falsely reporting new disease genes. Even though most of the new disease gene associations demonstrate multiple levels of evidence for the pathogenicity of discovered variants, a recent study by Hanany and colleagues demonstrated a high false discovery rate within autosomal dominant IRD genes that was a result of mistakenly classified pathogenic variants (Hanany and Sharon, 2019). In our case, both genes appeared to be strong candidates and only functional analyses led to the exclusion of both candidate genes as the cause of disease.

Re-analysis of sequencing data led to the detection of a causative 94 bp tandem duplication (NM_015629.4:c.73_166dup (p.Asp56fs) which creates a premature termination codon in *PRPF31*. The duplication was present in the unaffected mother and an unaffected sibling. The mother displayed no clinical manifestations of RCD even though she carried the mutation. The unaffected sibling was not studied clinically, however reported no symptoms. Additionally, she inherited an alternate wild type allele compared to her affected sibs further supporting partial penetrance that obscured the autosomal dominant inheritance pattern. In addition, the 94bp duplication is too large to be included in the standard high quality alignments and too small to be detected by read-depth CNV detection algorithms, such as gCNV (McKenna et al., 2010b). This led to the *PRPF31* variant being excluded in the initial variant filtering, even though loss of function variants in *PRPF31* are a known cause of dominant RCD through haploinsufficiency (Abu-Safieh et al., 2006). Structural changes have increasingly been shown to contribute to retinal disease, with an estimated contribution of CNVs of up to 9% of IRD cases (Ellingford et al., 2018; Van Cauwenbergh et al., 2017; Zampaglione et al., 2020). Gene size has been shown to be the strongest predictor for genes enriched with structural variation followed by long interspersed nuclear elements (LINE), long terminal repeats (LTR), and segmental duplications (Van Schil et al., 2018). For example, *USH2A* is one of the largest IRD genes (800.5 kb, hg19 chr1:215,796,236-216,596,738) and a recent publication found potentially causal structural variants in 9% of an Usher syndrome cohort (Bonnet et al., 2016b). *PRPF31*, however is an outlier. It is a small gene (16.3 kb, hg19 chr19:54,621,731-54,621,824) that is not in proximity to regions prone to non-homologous allelic recombination (NAHR)(Bujakowska et al., 2016), yet likely causal CNVs account for a large number of patients with *PRPF31*-associated disease (Sullivan et al., 2006; Zampaglione et al., 2020). *PRPF31* does contain a large number of total repeats relative to its size including low copy repeats that flank unique sequences which lead to non-recurrent CNVs (Carvalho and Lupski, 2016), as well as LINE and LTR (Van Schil et al., 2018). In the present case, the location of the duplication is in a low complexity region containing simple repeats, and the flanking sequences of the duplication contains a repeat (GCTATGGG) that may have facilitated DNA polymerase errors leading to the inclusion of the intervening region (Fig 3.E). The enrichment of genomic rearrangements combined with the complex inheritance pattern likely contributes to the underreporting of causal variants in *PRPF31*. Recent studies have shown diagnostic rates are improved by applying CNV analysis in an unbiased way in the initial analysis along with standard NGS pipelines, and this would allow for detection of causal CNVs in autosomal dominant genes such as *PRPF31* (Bonnet et al., 2016a; Ellingford et al., 2018). The overall contribution of *PRPF31* mutations in IRD is likely underestimated and should be considered in apparent autosomal recessive pedigrees without a clear solution in a known IRD gene.

This case study highlights the importance of thorough functional characterization of new candidate genes and the value of reanalysis of NGS sequence data as false discovery of novel genes can obscure true genetic diagnosis and may prevent the use of appropriate gene-based therapies in affected patients.

## METHODS

### Ethical guidelines

The study was approved by the institutional review board at the Massachusetts Eye and Ear (MEE), an affiliate of Mass General Brigham (MGB) healthcare system (Human Studies Committee MGB, Boston, MA, USA) and complied with the Health Information Portability and Accessibility Act (HIPAA). All aspects of the project adhered to the tenets of the Declaration of Helsinki. Informed consent was obtained from all individuals on whom genetic testing and further molecular evaluations were performed.

### Clinical evaluation

Ophthalmic evaluations were performed by clinicians experienced in inherited retinal degenerations at MEE. Visual acuity was measured using Snellen and Early Treatment of Diabetic Retinopathy Study (ETDRS) charts. Kinetic perimetry was performed using a Goldmann perimeter. Full-field electroretinograms (ERG) were performed using Burian Allen electrodes and custom ERG system with previously described parameters (Marmor et al., 2009; Reichel et al., 1989).

### Genetic screening

DNA extracted from venous blood using the DNeasy Blood and Tissue Kit (Qiagen, Hilden, Germany) was used for all the sequencing reactions. Whole exome sequencing (WES) was performed on OGI842-1649, OGI842-1651, OGI842-1653, OGI842-1831 and data processing were performed by the Genomics Platform at the Broad Institute of MIT and Harvard with an Illumina exome capture (38Mb target) and sequenced (150 bp paired reads) to cover >80% of targets at 20x and a mean target coverage of 100x. Exome sequencing data was processed through a pipeline based on Picard and mapping done using the BWA aligner to the human genome build 37 (hg19). The variant call sets were uploaded to variant analysis platform *seqr* (http://seqr.broadinstitute.org). Sanger sequencing was performed in all family members to validate potentially causal variants identified in WES including individuals who were not previously analyzed by WES (OGI842-1650, OGI842-1652).

### Splicing assay

Variants selected for analysis with the midigene assay were evaluated for possible effects on splicing using both Alamut software (Alamut Visual, http://www.interactive-biosoftware.com/alamut-visual/) and Splice AI neural network (Jaganathan et al., 2019). Two constructs containing *GNL3* exons 10-13 including intronic regions and *GNL3* with exons 13-15 along with downstream 3’ regions, were amplified from patient’s genomic DNA (Pfu Ultra II polymerase, Agilent Technologies, Santa Cruz, CA, primers listed in Table S1). The PCR products were cloned into pENTR-TOPO/D (ThermoFisher, Waltham, MA) and verified by Sanger sequencing. Clones carrying essential splice-site mutations were created by site-directed mutagenesis (QuickChange II Site Directed mutagenesis kit, Agilent Technologies, Santa Cruz, CA) on the wild-type constructs and verified by Sanger sequencing. Clones containing the desired sequences (wild-type, essential splice site and variant to be studied) were sub-cloned into pCSGW2+ vector (Jamshidi et al., 2019) by Gateway cloning using LR clonase (Invitrogen, Carlsbad, CA). pCSGW2+ *GNL3* constructs were transfected into HEK293 cells using 3ug of total DNA and Lipofectamine reagent (Invitrogen, Carlsbad, CA) into 6-well plates (Corning, Corning, NY) seeded with 5 × 10^5 cells. Human embryonic kidney cells (HEK293T) were grown on Dulbecco’s Modified Eagle’s Media (Gibco, Dublin, Ireland) and 10% fetal bovine serum (Thermo Fisher, Waltham, MA) at 37°C and 5% CO_2_. Total RNA was extracted from cells 72 hours after transfection (RNAeasy mini kit, QIAGEN, Hilden, Germany). cDNA was synthesized with Super Script II reverse transcriptase kit (Invitrogen, Carlsbad, CA) with 500ng RNA input. Finally, PCR amplification using a primer directed towards the plasmid and a second primer specific to the insert containing flanking exons of the splice-site mutations (Supplementary Table S1) was performed followed by visualization by agarose gel electrophoresis and NGS (Illumina, San Diego, CA).

STAR (version 2.5.3a) aligner (Dobin et al., 2012) was used to generate an index of the human genome (GRCh37.75.dna.primary_assembly.fa) and to align the reads for the analysis of splicing patterns from amplicon sequencing. Integrative Genomics Viewer (IGV) (Robinson et al., 2011) was used to load the aligned sequences (BAM files) for data visualization with Sashimi plots.

### Knockdown in Zebrafish

This study used an in-house cross of the AB and TL strain (source: ZIRC) for all experiments. Breeding and embryo maintenance were performed in accordance with MEE Zebrafish Core Facility standard procedures and IACUC approved protocols. Embryos were injected at the 1-8 cell stage, where each embryo received 2.3nL of either 0.4mM of the standard control MO or 0.2mM of each splice site blocking MO, (Supplementary Table S1). cDNA was extracted from each condition and analyzed via PCR and Sanger sequencing to confirm the effect on splicing. Zebrafish larvae external phenotypes were evaluated at day 5. Histology was performed by sectioning zebrafish larvae in paraffin blocks and Hematoxylin and Eosin staining ((SERI histology core, MGB).

To confirm morpholino results, CRISPR/Cas9 knockdown was performed targeting *pde4dip* exons 4 and 13. Zebrafish embryos were collected within 10 minutes of the beginning of breeding to ensure one-cell stage of the embryos. Both sgRNA (targeting exon 4 and 13, Supplementary Table S1) and Cas9 protein (EnGen Spy Cas9, NEB, Ipswich, MA) were co-injected into a cell cytoplasm of one-cell staged embryos. The mosaic F0 fish were kept for 5 days and analyzed functionally as morphant fish. Genomic DNA was extracted from the larvae after each assay and analyzed using NGS to observe cutting efficiency.

### Visual motor response and acoustically evoked behavior response

A combined visual motor response (VMR) and acoustically evoked behavior response (AEBR) assay was used to test the visual function and general motor function, respectively (Emran et al., 2008; Faria et al., 2019; Zeddies and Fay, 2005). In brief, larvae at 5 dpf were tested using the ZebraLab system (ViewPoint Life Sciences, Lyon, France). Larvae with body deformation or other severe external phenotypes were excluded. Individual larvae were placed in individual wells of a 96-well plate and placed in the Zebralab system, which is isolated from outside vibration and light. In the AEBR test, larvae were adapted for 10 min before being challenged by three consecutive vibrations (200 Hz for 500ms) with 5-min intervals under normal light illumination. In the VMR test, larvae were dark-adapted for 40 mins before going through three consecutive trials of light onset and light offset periods that last for 30 minutes each (Emran et al., 2008). The light change (on or off) was abrupt and was not fading. The animal’s motor response to the stimuli were recorded using infra-red light and camera. All the functional tests were performed between 10 am and 3 pm.

### Statistical analysis of zebrafish functional data

For the combined AEBR and VMR analysis, activity summarization and subsequent statistical analyses were performed using a customized analytical pipeline developed using R software (version 3.5.0, http://www.r-project.org/) building on previous analytical methods (Emran et al., 2008; Liu et al., 2015). The statistical analysis part of our analytical pipeline script is accessible in a GitHub repository here https://github.com/rongshisong/zebrafish_VMR_analysis. The differences between conditions and control in the mean activity level at the onset of the acoustic stimulation and during the first second after light change were tested using Tukey HSD with a p value of 0.05 or greater considered significant. All experiments were done with at least n=96 fish in each condition. Comparison of gas bladder presence was tested using Fisher’s exact test.

## Data Availability

All data produced in the present work are contained in the manuscript.

## ADDITIONAL INFORMATION

### Data deposition and Access

The causal variant identified in the present study has been deposited to ClinVar (https://www.ncbi.nih.gov/clinvar/) with accession number SCV001950339.

## Acknowledgements

This work was supported by grants from the National Eye Institute [R01EY026904 (KMB/EAP), R01EY012910 (EAP) and P30EY014104 (MEEI core support)], the Foundation Fighting Blindness (EGI-GE-1218-0753-UCSD, KMB) and the Curing Kids Foundation. Sequencing and analysis were provided by the Broad Institute of MIT and Harvard Center for Mendelian Genomics (Broad CMG) and was funded by the National Human Genome Research Institute, the National Eye Institute, and the National Heart, Lung and Blood Institute grant UM1HG008900 and in part by National Human Genome Research Institute grant R01 HG009141. The authors would like to thank the patients and their family members for their participation in this study and the Ocular Genomics Institute Genomics Core members for their experimental assistance. The authors would like to thank the Exome Aggregation Consortium, the Genome Aggregation Database (GnomAD) and the groups that provided exome variant data for comparison. A full list of contributing groups can be found at http://exac.broadinstitute.org/about and http://gnomad.broadinstitute.org/about.

## Author Contributions

H.S. and A.L. performed experiments and contributed to the writing of the manuscript. S.R. and S.M. provided data analysis. J.L.W. supported zebrafish data analysis and reviewed the manuscript. E.M.P. provided clinical data and reviewed the manuscript. E.A.P. contributed to the experimental design and review the manuscript. K.M.B. guided the experimental design, provided variant analysis and contributed to the writing of the manuscript.

## Notes

### Competing Interest Statement

The authors have declared no competing interest.

