## Supplemental Materials for "A Hidden Structural Variation in a Known IRD Gene: A Cautionary Tale of Two New Disease Candidate Genes"

**Table S1.**

| Primer name | Sequence |
|---|---|
| **Cloning** | |
| GNLS_exon11_splice_F | CACCTGACTGTGCTGAGTCTGTTC |
| GNLS_exon11_splice_R | TTTAACTCATGGAGGACACAC |
| GNLS_exon15_splice_F | CACCAATTGTGTGTCGCTGCTGTC |
| GNLS_exon15_splice_R | AGTTCAGCTAGCTGGTACAG |
| PRPF31_F | TGGCAGATGAGCTCTTAGCT |
| PRPF31_F | GGCTTGCTTGCTGATATACTCC |
| PRPF31_dup_F3 | TCGATCGCTGGCTCCTCTTCT |
| PRPF31_dup_R3 | GGAGGAGACACAGCTGGATCTT |
| **Zebrafish** | |
| Morpholino ex1_gnl3 | AGAATTAAACACTCACTCGGTCTCT |
| Morpholino TB_gnl3 | CAACTTCGGTCTCTTCATGTCTGCC |
| cDNA analysis F_gnl3 | GAAGTGAGGAGTTGAGACAC |
| cDNA analysis R_gnl3 | CAGCTTCTCGAAGCACTTCC |
| Morpholino-ex2_pde4dip | GCGCTACCAACTTTATACCTACCCA |
| Morpholino-ex18_pde4dip | TGCCGAATCCTGCGAGATTGATTGT |
| cDNA-ex2 analysis F_pde4dip | GATCAGAGCCACAGAAGAGAG |
| cDNA-ex2 analysis R_pde4dip | GAAGTTCTCCTTCTTCAGGTC |
| cDNA-ex18 analysis F_pde4dip | GCAGACAGACTCAAGAGCAT |
| cDNA-18 analysis R_pde4dip | GCTTCCTGACTTTCCACCTT |
| exon 4c sgRNA_pde4dip | GCGAGGATGTTCACCGCA**GGG** |
| exon 13 sgRNA_pde4dip | CCGTACAACGACACCGGG**AGG** |
| exon 4c GUIDE-IT F_pde4dip | AGAATCAAGTGCTACTGGTG |
| exon 4c GUIDE-IT R_pde4dip | GAGTGCAATAGTTAGTTCAC |
| exon 13 GUIDE-IT F_pde4dip | GAGCTTCTGTGCTCTCTAAA |
| exon 13 GUIDE-IT R_pde4dip | GCTCGTCTTACTTGTACCAG |
| exon 4c NGS F_pde4dip | CCTCAATGACCTGAAGAAGG |
| exon 4c NGS R_pde4dip | GTGGCTAATAACTGCCTGCC |
| exon 13 NGS F_pde4dip | CTTCAGAGGACACAGCAGCAG |
| exon 13 NGS R_pde4dip | CAGTCAGCAGGTCCTGGTTG |

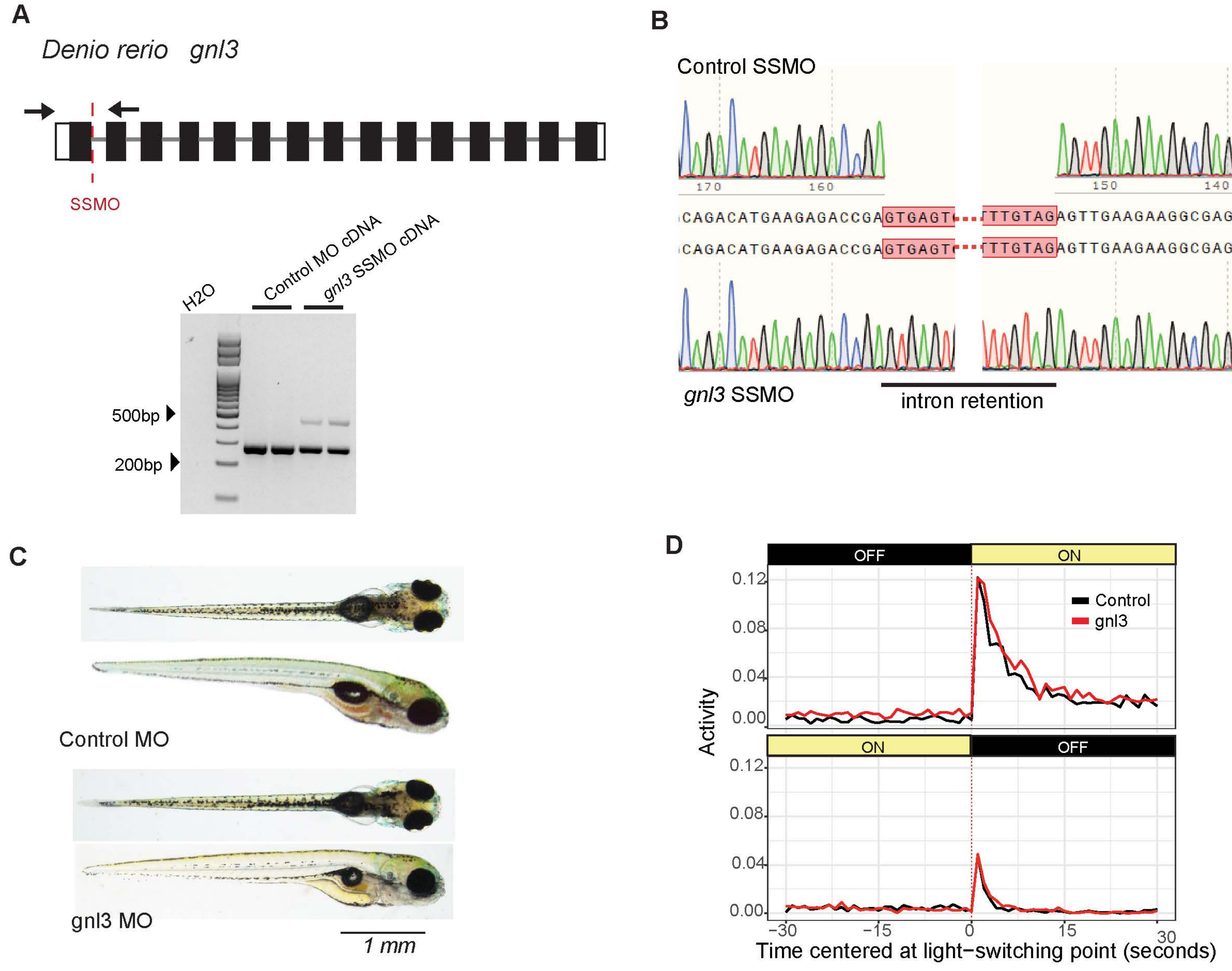


**Figure S1**. Functional studies in Zebrafish homolog *gnl3*. **(A)** Schematic of *D. rerio gnl3* with splice site morpholino (SSMO) targeting the exon/intron boundary of exon 1 indicated in a dashed red line and PCR primers indicated as black arrows. Below are results from RT_PCR performed on RNA isolated from control and *gnl3* morphant fish at 5 dpf, where upper band indicates intron retention in *gnl3* SSMO cDNA. **(B)** Chromatogram from Sanger sequencing of control and *gnl3* morphants, showing retained intron (black line and red boxes highlighting intronic sequences). **(C)** Zebrafish injected with control MO (top) and *gnl3* MO (bottom) at 5dpf with no observable differences in development or morphology. **(D)** Visual motor response (VMR) was performed on morphants at 5 dpf. The graph shows no significant difference between control MO (black, n=65) and *gnl3* MO knockdown (red, n=67) in either OFF/ON response (Tukey HSD, p=0.6352) or ON/OFF response (Tukey HSD p=0.8733).

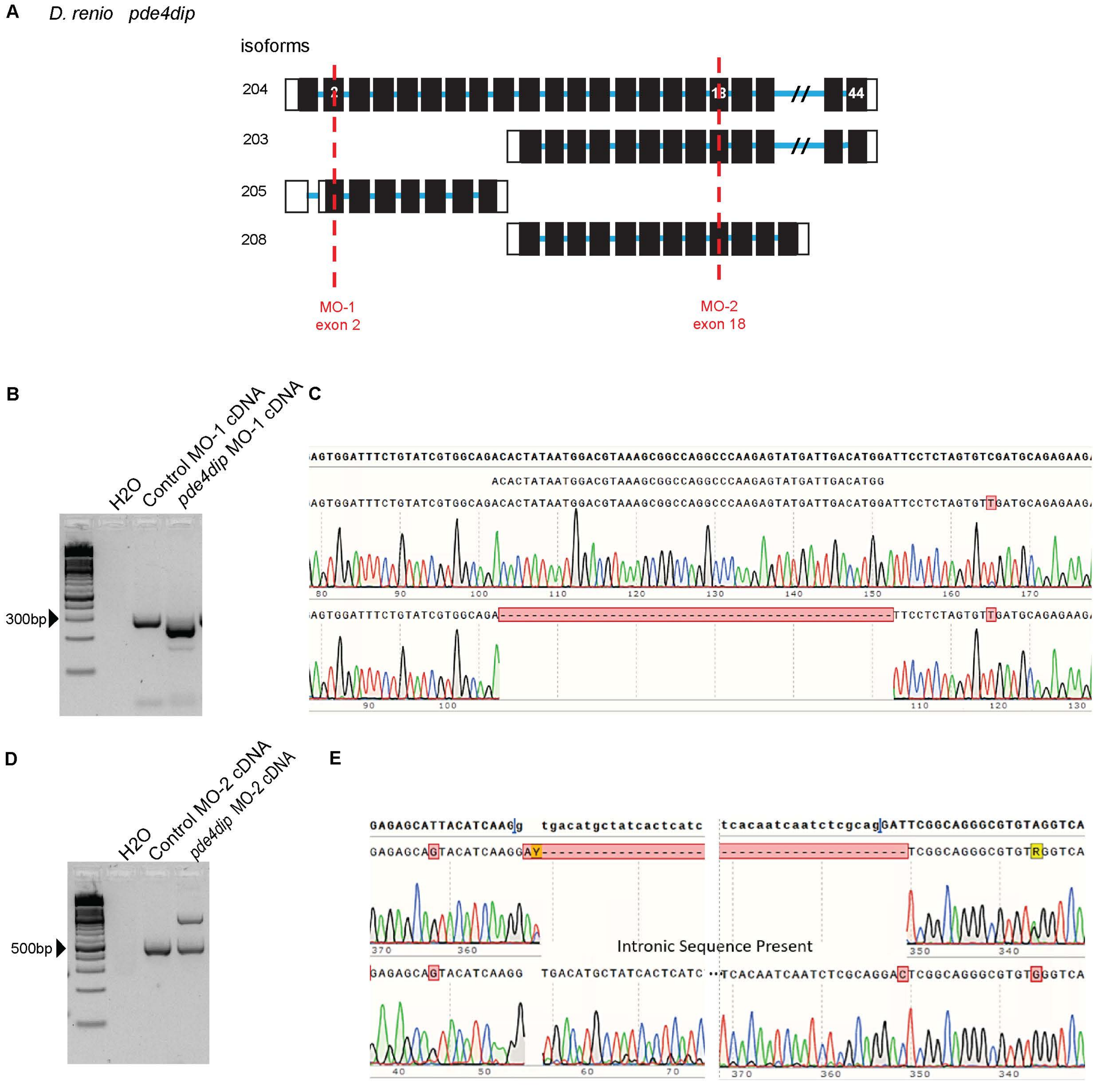


**Figure S2**. Functional studies in Zebrafish homolog *pde4dip*. **(A)** Schematic of *D. rerio pde4dip* depicts design of morpholinos to target exons 2 (MO-1) and 18 (MO-2) to ensure sufficient knock down of all isoforms. Morpholino targets are indicated with a dashed red line. **(B)** RT-PCR on RNA isolated from control and *pde4dip* morphant fish at 5 dpf to verify MO-1 efficiency. The last lane shows a lower band compared to control indicating exon skipping in pde4dip MO-1. **(C)** Sanger sequence demonstrating exon skipping due to MO-1. **(D)** RT-PCR on RNA isolated from control and *pde4dip* morphant fish at 5 dpf to verify MO-2 efficiency, where the upper band in the last lane indicates intron retention in *pde4dip* MO-2, with the lower band is similar to control MO. **(E)** Sanger sequence demonstrating intron retention due to MO-2.

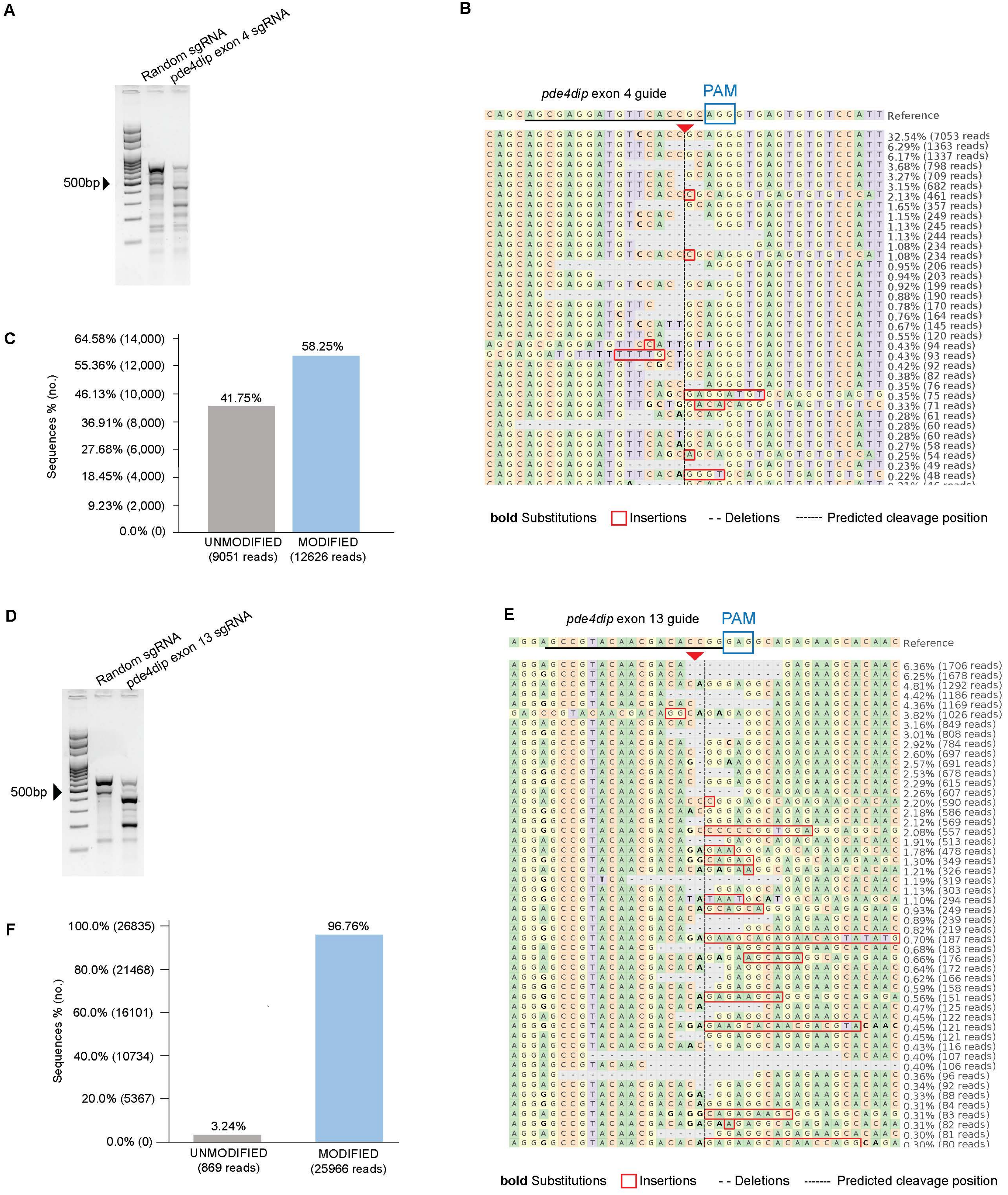


**Figure S3.** Validation of *pde4dip* CRISPR-cas9-editing in zebrafish. **(A)** T7E1 assay was performed on genomic DNA isolated from injected embryos to test for the editing efficiency of sgRNA exon 4 i*n vivo*. The last lane shows the depletion of the upper band compared to control, and lower bands indicate cutting in *pde4dip*-edited fish. **(B)** NGS analysis of genome editing of *pde4dip* exon 4 guide. Predicted cleavage site (dashed line) surrounded by editing events including substitutions (bold), insertions (red box), and deletions (- - -). Targeted sequence is underlined with protospacer adjacent motif (PAM) shown in blue box. Genomic DNA was isolated from random sgRNA control and pde4dip-edited fish at 5dpf. **(C)** Quantification of editing efficiencies show 58.25% of reads have been successfully modified. **(D)** T7E1 assay performed on genomic DNA to test editing efficiency. The last lane show lower bands compared to control that indicates cutting in *pde4dip* exon 13 sgRNA. **(E)** NGS analysis of genome editing of *pde4dip* exon 13 guide. Predicted cleavage site (dashed line) surrounded by editing events including substitutions (bold), insertions (red box), and deletions (- - -). Targeted sequence is underlined with protospacer adjacent motif (PAM) shown in blue box. Genomic DNA was isolated from random sgRNA control and pde4dip-edited fish at 5dpf. **(F)** Quantification of editing efficiencies show 96.76% of reads have been successfully modified. (CRISPResso2 http://crispresso.pinellolab.partner.org/)

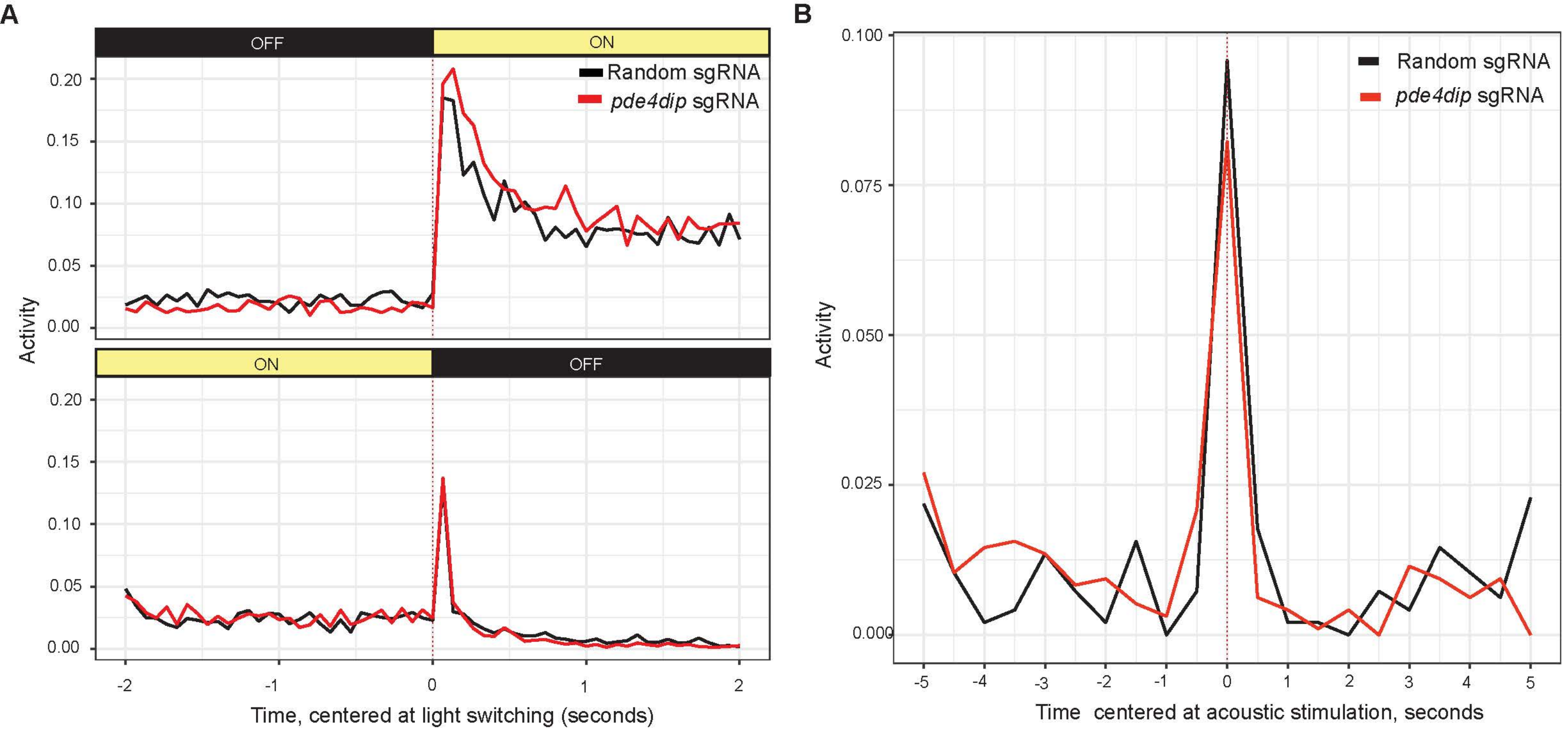


**Figure S4.** **(A)** VMR assay was performed on *pde4dip* CRISPR-Cas9 edited fish at 5 dpf. The graph shows no significant difference between random sgRNA (black, n=48) and *pde4dip* sgRNA knockdown (red, n=48) in either OFF/ON response (t-test, p=0.8079) or ON/OFF response (t-test, p=0.5417). **(B)** AEBR assay performed on *pde4dip* CRISPR-Cas9 edited fish at 5 dpf shows no significant difference in response to stimuli between *pde4dip* sgRNA (red, n=32) and fish injected with random sgRNA (black, n=32) (t-test, p=0.6465).
